# Older adults mount less durable humoral responses to two doses of COVID-19 mRNA vaccine, but strong initial responses to a third dose

**DOI:** 10.1101/2022.01.06.22268745

**Authors:** Francis Mwimanzi, Hope R. Lapointe, Peter K. Cheung, Yurou Sang, Fatima Yaseen, Gisele Umviligihozo, Rebecca Kalikawe, Sneha Datwani, F. Harrison Omondi, Laura Burns, Landon Young, Victor Leung, Olga Agafitei, Siobhan Ennis, Winnie Dong, Simran Basra, Li Yi Lim, Kurtis Ng, Ralph Pantophlet, Chanson J. Brumme, Julio S.G. Montaner, Natalie Prystajecky, Christopher F. Lowe, Mari L. DeMarco, Daniel T. Holmes, Janet Simons, Masahiro Niikura, Marc G. Romney, Zabrina L. Brumme, Mark A. Brockman

**Author notes:** **Corresponding Author Contact Information:** Mark A. Brockman, Ph.D., Professor, Faculty of Health Sciences, Simon Fraser University, 8888 University Drive, Burnaby, BC, Canada, V5A 1S6. MAB, MGR and ZLB contributed equally.

## Abstract

**Background:** Third COVID-19 vaccine doses are broadly recommended, but immunogenicity data remain limited, particularly in older adults.

**Methods:** We measured circulating antibodies against the SARS-CoV-2 spike protein receptor-binding domain, ACE2 displacement, and virus neutralization against ancestral and Omicron (BA.1) strains from pre-vaccine up to one month following the third dose, in 151 adults aged 24-98 years who received COVID-19 mRNA vaccines.

**Results:** Following two vaccine doses, humoral immunity was weaker, less functional and less durable in older adults, where a higher number of chronic health conditions was a key correlate of weaker responses and poorer durability. Third doses boosted antibody binding and function to higher levels than second-doses, and induced responses in older adults that were comparable in magnitude to those in younger adults. Humoral responses against Omicron were universally weaker than against the ancestral strain after both second and third doses; nevertheless, after three doses, anti-Omicron responses in older adults reached equivalence to those in younger adults. After three vaccine doses, the number of chronic health conditions, but not age per se, was the strongest consistent correlate of weaker humoral responses.

**Conclusion:** Results underscore the immune benefits of third COVID-19 vaccine doses, particularly in older adults.

## INTRODUCTION

Older adults are at increased risk of lethal COVID-19 following SARS-CoV-2 infection (SARS-CoV-2) [1-3]. While two doses of a COVID-19 mRNA vaccine broadly protects against hospitalization and death [4-6], weaker vaccine-induced immunity observed in the elderly and certain other groups [7-12] has led to their prioritization to receive third doses [13-16]. Vaccine-induced antibodies also decline over time, which can increase the risk of breakthrough infections [17-19], particularly with the more transmissible and immune evasive Omicron variant (B.1.1.529) [20-22].

We and others have shown that older age is associated with weaker antibody responses to COVID-19 mRNA vaccines, Comirnaty (Pfizer/BioNTech) and Spikevax (Moderna) [10-12]. We previously characterized longitudinal humoral responses up to three months after the second vaccine dose in a cohort of 151 adults 24 to 98 years of age that includes COVID-19 naïve and convalescent individuals [12]. Here, we examine binding and neutralizing antibody responses up to six months following the second vaccine dose, as well as one month following the third vaccine dose. We also evaluate binding antibodies, ACE2 displacement, and virus neutralization against Omicron (BA.1) one month following the second and third doses.

## METHODS

### Study design

We conducted a prospective longitudinal cohort study in British Columbia, Canada, to examine SARS-CoV-2 specific humoral responses following vaccination with Comirnaty or Spikevax. Our cohort of 151 individuals included 81 healthcare workers (HCW) and 56 older adults (including 18 residents of long-term care or assisted living facilities) who were COVID-19 naive at study entry, and 14 COVID-19 convalescent individuals with anti-SARS-CoV-2 N antibodies at study entry (including 8 HCW and 6 older adults) [12]. Serum and plasma were collected prior to vaccination; one month after the first dose; one, three and six months after the second dose; and one month following the third dose. Specimens were processed same-day and frozen until analysis.

### Ethics approval

Written informed consent was obtained from all participants or their authorized decision makers. This study was approved by the University of British Columbia/Providence Health Care and Simon Fraser University Research Ethics Boards.

### Data sources

Sociodemographic, health and vaccine information was collected by self-report and confirmed through medical records where available. Chronic health conditions were defined as hypertension, diabetes, asthma, obesity (body mass index ≥30), chronic diseases of lung, liver, kidney, heart or blood, cancer, and immunosuppression due to chronic conditions or medication, to generate a score ranging from 0-11 per participant [12].

### Binding antibody assays

We measured total binding antibodies against SARS-CoV-2 nucleocapsid (N) and spike (S) receptor binding domain (RBD) in serum using the Roche Elecsys Anti-SARS-CoV-2 and Anti-SARS-CoV-2 S assays, respectively, on a Cobas e601 module analyzer (Roche Diagnostics). Following SARS-CoV-2 infection, both assays should be positive, whereas post-vaccination only the S assay should be positive, allowing identification of convalescent individuals. Both tests are electro-chemiluminescence sandwich immunoassays, and report results in Arbitrary Units (AU)/mL, calibrated against an external standard. For the S assay, the manufacturer indicates that AU values can be considered equivalent to international binding antibody units (BAU) as defined by the World Health Organization [23]. For the S assay, sera were tested undiluted, with samples above the upper limit of quantification (ULOQ) re-tested at 1:100 dilution, allowing a measurement range of .0.4 - 25,000 U/mL. We also quantified plasma IgG binding antibodies against RBD using the V-plex SARS-CoV-2 (IgG) Panel 22 ELISA kit (Meso Scale Diagnostics), which features the ancestral (Wuhan) and Omicron RBD antigens, on a Meso QuickPlex SQ120 instrument. Plasma samples were diluted 1:10000 as directed by the manufacturer, with results reported in Arbitrary Units (AU)/mL.

### ACE2 competition assay

We assessed the ability of plasma antibodies to block the RBD-ACE2 receptor interaction by competition ELISA (Panel 22 V-plex SARS-CoV-2 [ACE2]; Meso Scale Diagnostics) on a Meso QuickPlex SQ120 instrument. Plasma was diluted 1:20 as directed by the manufacturer and results reported as % ACE2 displacement.

### Live virus neutralization

Neutralizing activity in plasma was examined using a live SARS-CoV-2 infectivity assay in a Containment Level 3 facility. Assays were performed using isolate USA-WA1/2020 (BEI Resources) and a local Omicron isolate (BA.1 strain; GISAID Accession # EPI_ISL_9805779) on VeroE6-TMPRSS2 (JCRB-1819) target cells. Viral stock was adjusted to 50 TCID_50_/200 µl in Dulbecco’s Modified Eagle Medium in the presence of serial 2-fold dilutions of plasma (from 1/20 to 1/2560), incubated at 4°C for 1 hour and then added to target cells in 96-well plates in triplicate. Cultures were maintained at 37°C with 5% CO_2_ and the appearance of viral cytopathic effect (CPE) was recorded three days post-infection. Neutralizing activity is reported as the highest reciprocal plasma dilution able to prevent CPE in all three replicate wells. Samples exhibiting only partial or no neutralization at the lowest dilution of 1/20 were coded as having a reciprocal dilution of “10”, defined as below the limit of quantification (BLOQ) in this assay.

### Statistical analysis

Comparisons of binary variables were performed using Fisher’s exact test. Comparisons of continuous variables were performed using the Mann-Whitney U-test (for unpaired data) or Wilcoxon test (for paired data). Multiple linear regression was used to investigate the relationship between sociodemographic, health and vaccine-related variables and humoral outcomes. Variables included age (per year increment), sex at birth (female as reference group), ethnicity (non-white as reference), number of chronic health conditions (per number increment), mRNA vaccine received (Comirnaty as reference), interval between doses (per day increment), sampling date following the most recent dose (per day increment), and convalescent status (COVID-19 naive as reference). Binding antibody half-lives in serum were calculated by fitting exponential decay curves to antibody concentrations at one, three and six months after the second dose. All tests were two-tailed, with p<0.05 considered statistically significant. Analyses were conducted using Microsoft Excel and Prism v9.2.0 (GraphPad).

## RESULTS

### Participant characteristics

As described previously [12], the cohort is predominantly female (**Table 1**). HCW, older adults and COVID-19 convalescent individuals at study entry were a median of 41, 79 and 48 years old, respectively. Older adults were predominantly (77%) of white ethnicity (compared to 46% of HCW) and had a higher burden of chronic health conditions (a median of 1, interquartile range [IQR] 0-2, range 0-5, vs. a median of 1, IQR 0-0, range 0-3 in HCW). All participants received two COVID-19 mRNA vaccine doses between December 2020-July 2021, where the dose interval was up to 112 days as per national guidelines to delay second doses due to initially limited vaccine supply. A total of 141 (93%) and 138 (91%) of participants received Comirnaty as their first and second dose, respectively. At the time of writing, 114 participants had received a third dose between October-December 2021, on average 7 months following their second dose. For participants whose third dose was Spikevax (53% of the cohort), those aged ≥70 years received a full dose, whereas those <70 years received a half-dose, as per national guidelines. An additional six (7.4%) HCW and two (3.6%) older adults developed anti-N antibodies during follow-up, reflecting breakthrough infections. Three of these infections, all in HCW, occurred between December 2021-Jan 2022 and are likely Omicron. In longitudinal analyses that span the entire study, participants with a post-vaccination SARS-CoV-2 infection are retained in their original “COVID-19 naive at study entry” groups but identified in the Figures, while in analyses that focus on third dose responses, they are grouped in a single “prior COVID-19” group.

**Table 1:** Participant characteristics and sampling information.

| Variable category | Characteristic | Healthcare Workers (n=81) | Older Adults (n=56) | COVID-19 Convalescent at study entry (n=14) |
| --- | --- | --- | --- | --- |
| Sociodemographic/health | Age in years, median [IQR]<sup>a</sup> | 41 [35-51] | 78 [73-83] | 48 [36-87] |
|  | Female sex, n (%) | 61 (75%) | 38 (68%) | 10 (71%) |
|  | White/Caucasian ethnicity, n (%) | 37 (46%) | 43 (77%) | 7 (50%) |
|  | Chronic health or immunosuppressive conditions, median [IQR] | 0 [0-0] | 1 [0-2] | 0 [0-1] |
| Vaccine information | Comirnaty, First mRNA Vaccine, n (%) | 80 (99%) | 48 (86%) | 13 (93%) |
|  | Comirnaty, Second mRNA Vaccine, n (%) | 79 (98%) | 46 (82%) | 13 (93%) |
|  | Time between first and second doses in days, median [IQR] | 97 [91-102] | 76 [45-85] | 112 [87-118] |
|  | Comirnaty, Third mRNA Vaccine, n (%)<sup>b</sup> | 32/61 (52%) | 19/47 (40%) | 3/6 (50%) |
|  | Time between second and third dose in days, median [IQR] | 210 [200-241] | 169 [160-231] | 189 [170-194] |
| Specimen collection | Specimens collected pre-vaccine, n (%) | 80 (99%) | 49 (88%) | 13 (93%) |
|  | Specimens collected one month after first dose, n (%) | 79 (98%) | 49 (88%) | 13 (93%) |
|  | Day of specimen collection one month after first dose, median [IQR] days | 28 [27-30] | 30 [28-32] | 31 [28-32] |
|  | Specimens collected one month after second dose, n (%) | 81 (100%) | 55 (98%) | 14 (100%) |
|  | Day of specimen collection one month after second dose, median [IQR] days | 29 [29-32] | 29 [29-31] | 32 [30-36] |
|  | Specimens collected three months after second dose, n (%) | 79 (98%) | 53 (95%) | 13 (93%) |
|  | Day of specimen collection three months after second dose, median [IQR] days | 90 [90-91] | 90 [89-92] | 90 [87-91] |
|  | Specimens collected six months after second dose, n (%) | 78 (96%) | 40 (71%) | 10 (71%) |
|  | Day of specimen collection six months after second dose, median [IQR] days | 181 [179-182] | 176 [167-182] | 180 [179-181] |
|  | Specimens collected one month after third dose, n (%) | 61 (75%) | 47 (84%) | 6 (38%) |
|  | Day of specimen collection one month after third dose, median [IQR] days | 30 [29-31] | 32 [29-33] | 30 [29-30] |
| COVID-19 post-vax | Anti-N seroconversion during study follow-up | 6 (7.4%) | 2 (3.6%) | - |

<sup>a</sup> interquartile range<sup>b</sup> denominators are the n of specimens collected one month after third dose

### After two-dose vaccination, lower binding antibodies are associated with older age and burden of chronic health conditions, but older adults mount strong responses after a third dose

We measured total anti-RBD binding antibody concentrations in serum before and after immunization (**Figure 1A)**. As reported previously [12], antibody concentrations in older adults were significantly lower than those in HCW one month after the first dose (a median of 2.00 [IQR 1.75-2.25] log_10_ U/mL in HCW versus a median of 1.50 [IQR 1.05-1.99] in older adults), as well as one month after the second dose (a median of 4.02 [IQR 3.88-4.25] in HCW versus a median of 3.74 [IQR 3.49-3.91] in older adults) (Mann-Whitney; both p<0.0001). Three months following the second dose, antibody concentrations had declined by ∼0.4 log_10_ on average, to a median of 3.63 [IQR 3.44-3.83] in HCW versus a median 3.32 [IQR 3.04-3.56] in older adults) (Mann-Whitney p<0.0001 for comparison between groups). Six months following the second dose, antibody concentrations had declined by a further ∼0.3 log_10_ on average, to a median of 3.30 [IQR 3.09-3.47] in HCW versus a median 2.96 [IQR 2.68-3.20] in older adults (p<0.0001). This confirms that, following two-dose COVID-19 mRNA vaccination, antibody concentrations remain consistently and significantly lower in older compared to younger adults. By contrast, antibody concentrations in COVID-19 convalescent individuals remained consistently higher than COVID-19 naive individuals at all time points after two doses. Six months after the second dose for example, convalescent individuals maintained median responses of 3.50 (IQR 3.40-3.71) log_10_ U/mL (p=0.027 compared to HCW; p<0.0001 compared to older adults).

**Figure 1.**
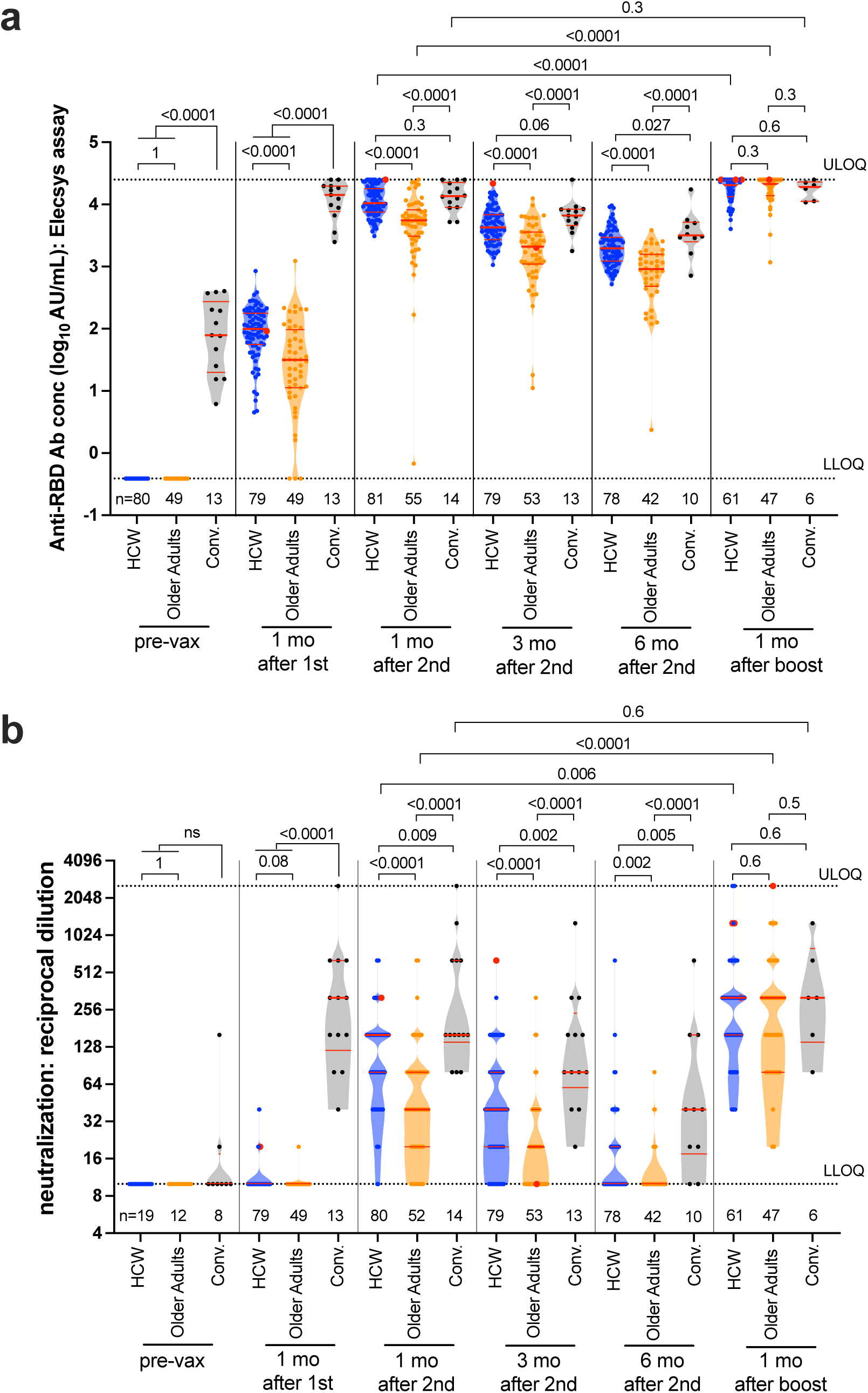
Longitudinal antibody binding and neutralization responses to spike RBD following one, two and three COVID-19 vaccine doses. *Panel A:* Binding antibody responses to the SARS-CoV-2 spike RBD in serum, in HCW (blue circles) and older adults (orange circles) who were COVID-19 naive at study entry, as well as COVID-19 convalescent individuals (black circles) at six timepoints: prior to vaccination (pre-vax), one month following the first dose, one, three and six months following the second dose, and one month following the third vaccine dose. Individuals with post-vaccination infections are indicated by red dots at their first N seropositive time point. Participant Ns are provided at the bottom of the plot. A thick horizontal red bar represents the median; thinner horizontal red bars represent the IQR. P-values were computed using the Mann-Whitney U-test (for comparisons between groups) or the Wilcoxon matched pairs test (for comparisons across time points within a group) and are uncorrected for multiple comparisons. ULOQ/LLOQ: upper/lower limit of quantification. *Panel B:* same as A, but for virus neutralization activity, defined as the lowest reciprocal plasma dilution at which neutralization was observed in all wells of a triplicate assay. Plasma samples showing neutralization in fewer than three wells at a 1/20 dilution were coded as having a reciprocal dilution of 10, corresponding to the LLOQ in this assay. The highest dilution tested was 1/2560, which corresponds to the ULOQ. Note that only a subset of pre-vaccine plasma samples was assayed for this activity.

Multivariable analyses of antibody concentrations after two doses, that adjusted for sex, ethnicity, number of chronic health conditions, first-dose vaccine brand, dosing interval and day of specimen collection post-immunization confirmed that older age remained independently associated with lower antibody concentrations at one and three months after the second dose (**Table S1**). One month following the second dose for example, each decade of older age was associated with an ∼0.06 log_10_ lower antibody concentration (p=0.0067). A higher number of chronic conditions was also independently associated with lower antibody concentrations at both these time points. Six months following the second dose, a higher number of chronic health conditions remained the strongest independent correlate of lower responses, with each additional condition associated with an 0.14 log_10_ lower antibody concentration (p=0.0001). A longer dose interval was also associated with higher antibody concentrations at all time points after the second dose (all p<0.05), consistent with previous reports [24-26]. COVID-19 convalescent status was also associated with maintaining 0.26 log_10_ higher antibody concentrations at three and six months following the second dose (both p<0.05), consistent with superior durability of “hybrid” immunity induced by infection followed by vaccination [27-29].

In both HCW and older adults, the third dose boosted antibody concentrations at least ∼0.3-0.4 log_10_ higher than peak values observed after two doses (Wilcoxon paired test p<0.0001 for both groups). Binding antibodies in HCW rose to a median of 4.31 (IQR 4.13 to upper limit of quantification [ULOQ]) whereas those in older adults rose to a median of 4.33 (4.14 to ULOQ) (p=0.33), indicating that older and younger adults mounted comparable initial binding antibody responses following a third dose. In multivariable analyses of third-dose responses, a higher number of chronic health conditions was the sole significant correlate of lower antibody concentrations (p=0.0078), while having received Spikevax as the third dose was associated with higher antibody concentrations (p=0.0091) (**Table S2**).

### After two-dose vaccination, weaker virus neutralizing activity is associated with age and chronic health conditions, but older adults mount strong responses after a third dose

We performed live SARS-CoV-2 neutralization assays to quantify the ability of plasma to block virus infection of target cells (**Figure 1B**). Neutralizing activity is reported as the highest reciprocal plasma dilution capable of preventing viral cytopathic effects in all wells of a triplicate assay, where a reciprocal dilution of “10” indicates no or limited neutralization. As previously reported [12], one vaccine dose largely failed to induce neutralizing activity in COVID-19 naïve individuals, though two doses induced this activity in most participants, albeit at consistently lower levels in older compared to younger adults. One month after the second dose for example, the median reciprocal dilution was 160 [IQR 80-160] in HCW versus 40 [IQR 20-80] in older adults (p<0.0001). Three months after the second dose, neutralizing activity had declined by more than two-fold on average, to a median reciprocal dilution of 40 (IQR 20-80) in HCW versus a median of 20 (IQR BLOQ-40) in older adults (p<0.0001). Six months after the second dose, neutralizing activity had declined to below the limit of quantification (BLOQ) in 58% of HCW and 83% of older adults (Mann-Whitney p=0.0048 for comparison between groups). COVID-19 convalescent individuals by contrast maintained significantly higher neutralizing activity compared to naive individuals at all time points following two-dose vaccination. Multivariable analyses confirmed that older age remained significantly associated with weaker neutralizing activity at one and three months after two-dose vaccination, while COVID-19 convalescent status was associated with superior neutralizing activity at all time points following two-dose vaccination (all p≤0.0002) (**Table S1**).

A third vaccine dose boosted neutralizing activity in both HCW and older adults, achieving responses that were two-fold and eight-fold higher than peak values after two doses, respectively (Wilcoxon paired test p≤0.006 for both groups; **Figure 1B**). Specifically, the median reciprocal dilution in HCW and older adults rose to 320 [IQR 160-320] and 320 [IQR 80-320], respectively (p=0.6), indicating that older adults mounted comparable neutralizing responses to younger adults after three doses. A multivariable analysis identified prior COVID-19 as the strongest independent predictor of higher neutralizing activity after a third vaccine dose (p=0.0044; **Table S2**).

### After two-dose vaccination, binding antibody responses decline faster in those with a higher burden of chronic conditions

We next assessed temporal reductions in antibody concentrations after two-dose vaccination (**Figure 2A**). Assuming exponential decay and restricting the analysis to participants with a complete longitudinal data series with no values above the ULOQ, we estimated antibody concentration half-lives to be a median of 59 [IQR 52-75] days in HCW versus a median of 52 [IQR 45-65] days in older adults (p=0.016; **Figure 2B**). This suggests that, in addition to mounting overall weaker responses to two-dose vaccination compared to younger adults, antibody concentrations in older adults also decline more rapidly. In multivariable analyses however, a higher number of chronic health conditions emerged as the sole independent correlate of antibody decline, with each additional condition associated with a 5-day shorter half-life (p=0.017; **Table 2**). Furthermore, COVID-19 convalescent status was associated with a 14-day longer antibody half-life after adjustment for other factors (p=0.056), consistent with improved durability of hybrid immunity [27-29].

**Table 2:** Multivariable analysis of the relationship between sociodemographic, health and vaccine-related variables on serum antibody half-life following two-dose COVID-19 mRNA vaccination.

| **Outcome measure** | **Variable** | **Estimate** | **95% CI** | **p-value** |
| --- | --- | --- | --- | --- |
| Ab half-life after two vaccine doses | Age (per year) | 0.058 | -0.17 to 0.29 | 0.61 |
|  | Male sex | 5.31 | -2.57 to 13.18 | 0.18 |
|  | White ethnicity | 3.11 | -4.67 to 10.88 | 0.43 |
|  | # chronic conditions (per add'l) | -4.62 | -8.39 to -0.85 | **0.017** |
|  | Spikevax as first dose | 3.37 | -13.26 to 20.00 | 0.69 |
|  | Dose interval (per day) | 0.00014 | -0.16 to 0.16 | 0.99 |
|  | COVID-19 convalescent<sup>a</sup> | 13.78 | -0.37 to 27.93 | 0.056 |

<sup>a</sup> participants with positive anti-N serology at study entry

**Figure 2:**
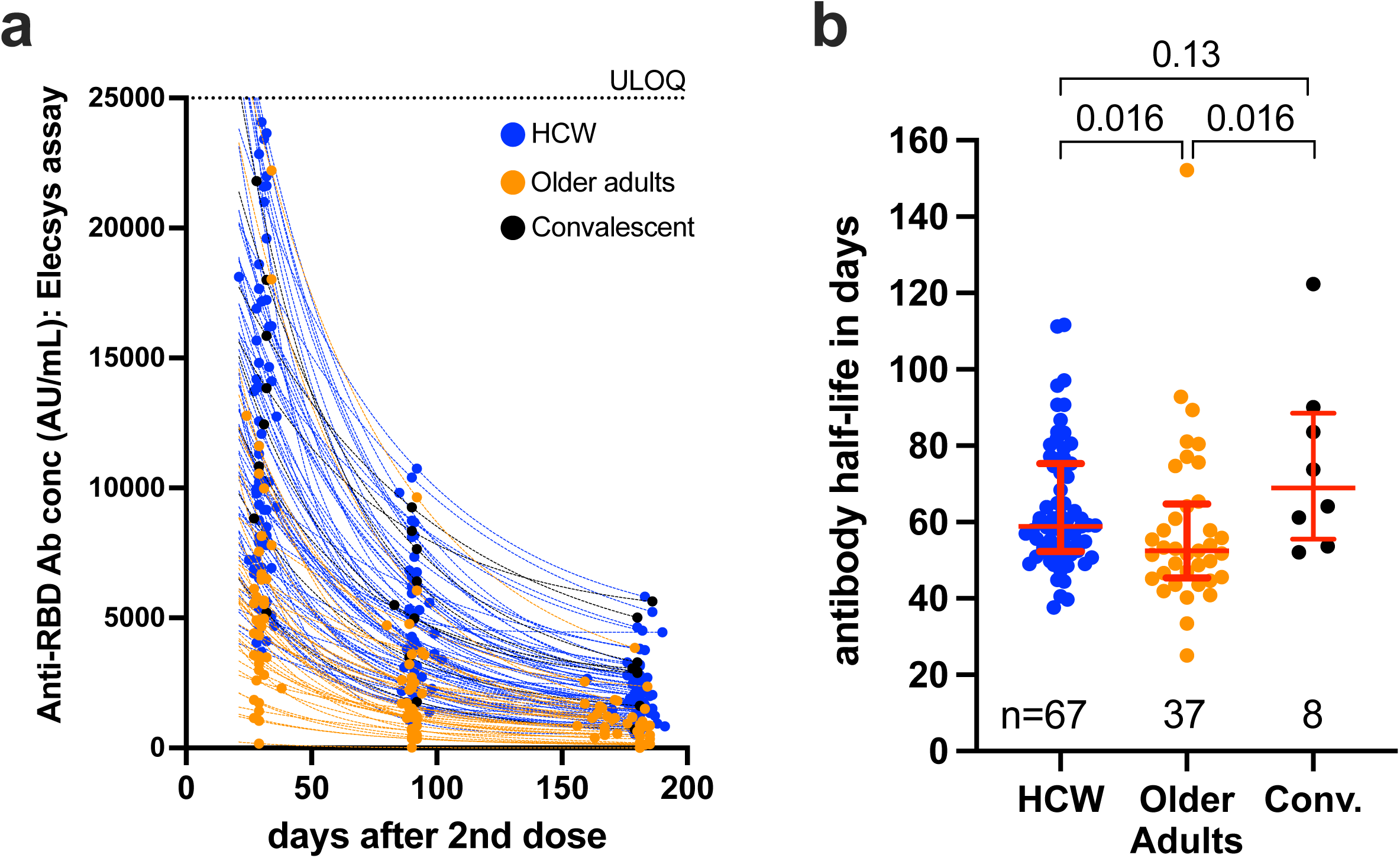
Decay rates of serum binding antibody responses to spike RBD following two COVID-19 vaccine doses. *Panel A:* Temporal declines in serum binding antibody responses to spike RBD following two vaccine doses in HCW (blue) and older adults (orange) who were COVID-19 naive at study entry, as well as COVID-19 convalescent participants (black circles). ULOQ: upper limit of quantification. Only participants with a complete longitudinal data series with no values above the ULOQ are shown. *Panel B*: Binding antibody half-lives following two COVID-19 vaccine doses, calculated by fitting an exponential curve to each participant’s data shown in panel A. Participant Ns are indicated at the bottom of the plot. Red bars and whiskers represent the median and IQR. P-values were computed using the Mann-Whitney U-test and are uncorrected for multiple comparisons.

### Humoral responses against Omicron following two and three vaccine doses

Given the rapid rise of the Omicron variant, we compared peak antibody responses against this strain in plasma collected at one month after the second and third vaccine doses. Here, we grouped all participants with prior COVID-19, regardless of infection timing, in the convalescent category. Overall, IgG binding antibodies against the Omicron RBD, measured using the Meso Scale Diagnostics V-Plex assay, were on average 0.4 to 0.5 log_10_ U/mL lower than those against the wild type (WT; ancestral Wuhan strain) RBD antigen after two and three doses (all within-group comparisons p≤0.0002; **Figure 3A**). Nevertheless, the third dose universally boosted anti-Omicron IgG concentrations to an average of 0.5 log_10_ higher than levels induced by two doses (all within-group comparisons p<0.05). Consistent with total binding antibody concentrations quantified using the Roche assay (**Figure 1A**), binding IgG concentrations against the WT RBD were significantly higher in HCW compared to older adults after two doses (p<0.0001) but reached equivalence after three doses (p=0.4). IgG concentrations capable of binding Omicron followed a similar pattern, with HCW showing marginally higher anti-Omicron IgG levels compared to older adults after two doses (p=0.09), but equivalent levels after three doses (p=0.49). A multivariable analysis of Omicron-specific IgG concentrations after three doses identified a higher number of chronic health conditions as the strongest correlate of poorer responses, with each additional condition associated with a 0.12 log_10_ reduction in Omicron binding IgG (p=0.0033; **Table 3**). A longer interval between the first and second vaccine doses was marginally associated with a lower third dose response (p=0.02).

**Table 3:** Multivariable analyses of the relationship between sociodemographic, health and vaccine-related variables on Omicron-specific humoral immunogenicity measures following three-dose COVID-19 mRNA vaccination.

| Humoral measure | Variable | 1 mo after 3rd dose |  |  |
| --- | --- | --- | --- | --- |
|  |  | Estimate | 95% CI | p-value |
| anti-Omicron RBD IgG (log10)<sup>a</sup> | Age (per year) | 0.0035 | -0.0027 to 0.0097 | 0.26 |
|  | Male sex | -0.14 | -0.34 to 0.054 | 0.15 |
|  | White ethnicity | -0.018 | -0.21 to 0.17 | 0.85 |
|  | # chronic conditions (per add'l) | -0.12 | -0.20 to -0.041 | **0.0033** |
|  | Spikevax as third dose (vs. Comirnaty) | 0.15 | -0.039 to 0.34 | 0.12 |
|  | Interval between 1st and 2nd dose (per day) | -0.0066 | -0.012 to -0.0011 | **0.020** |
|  | Interval between 2nd and 3rd dose (per day) | 0.00043 | -0.0034 to 0.0042 | 0.83 |
|  | Days since 3rd vaccine dose | -0.0086 | -0.038 to 0.021 | 0.56 |
|  | Prior COVID-19<sup>b</sup> | 0.1 | -0.16 to 0.37 | 0.43 |
| anti-Omicron ACE2 % displacement<sup>a</sup> | Age (per year) | 0.29 | -0.046 to 0.63 | 0.090 |
|  | Male sex | -12.38 | -23.16 to -1.60 | **0.025** |
|  | White Ethnicity | -2.36 | -12.74 to 8.01 | 0.65 |
|  | # chronic conditions (per add'l) | -6.41 | -10.80 to -2.03 | **0.0046** |
|  | Spikevax as third dose (vs. Comirnaty) | 1.69 | -8.58 to 11.97 | 0.74 |
|  | Interval between 1st and 2nd dose (per day) | -0.41 | -0.71 to -0.11 | **0.0079** |
|  | Interval between 2nd and 3rd dose (per day) | -0.038 | -0.25 to 0.17 | 0.72 |
|  | Days since 3rd vaccine dose | -1.82 | -3.43 to -0.21 | **0.027** |
|  | Prior COVID-19<sup>b</sup> | 12.28 | -2.11 to 26.67 | 0.094 |

<sup>a</sup> Measured using the Meso Scale Diagnostics (MSD) V-plex assay system
<sup>b</sup> Includes all participants with positive anti-N serology at any time during the study (*i.e.* both pre- and post-vaccine COVID-19 cases)

**Figure 3:**
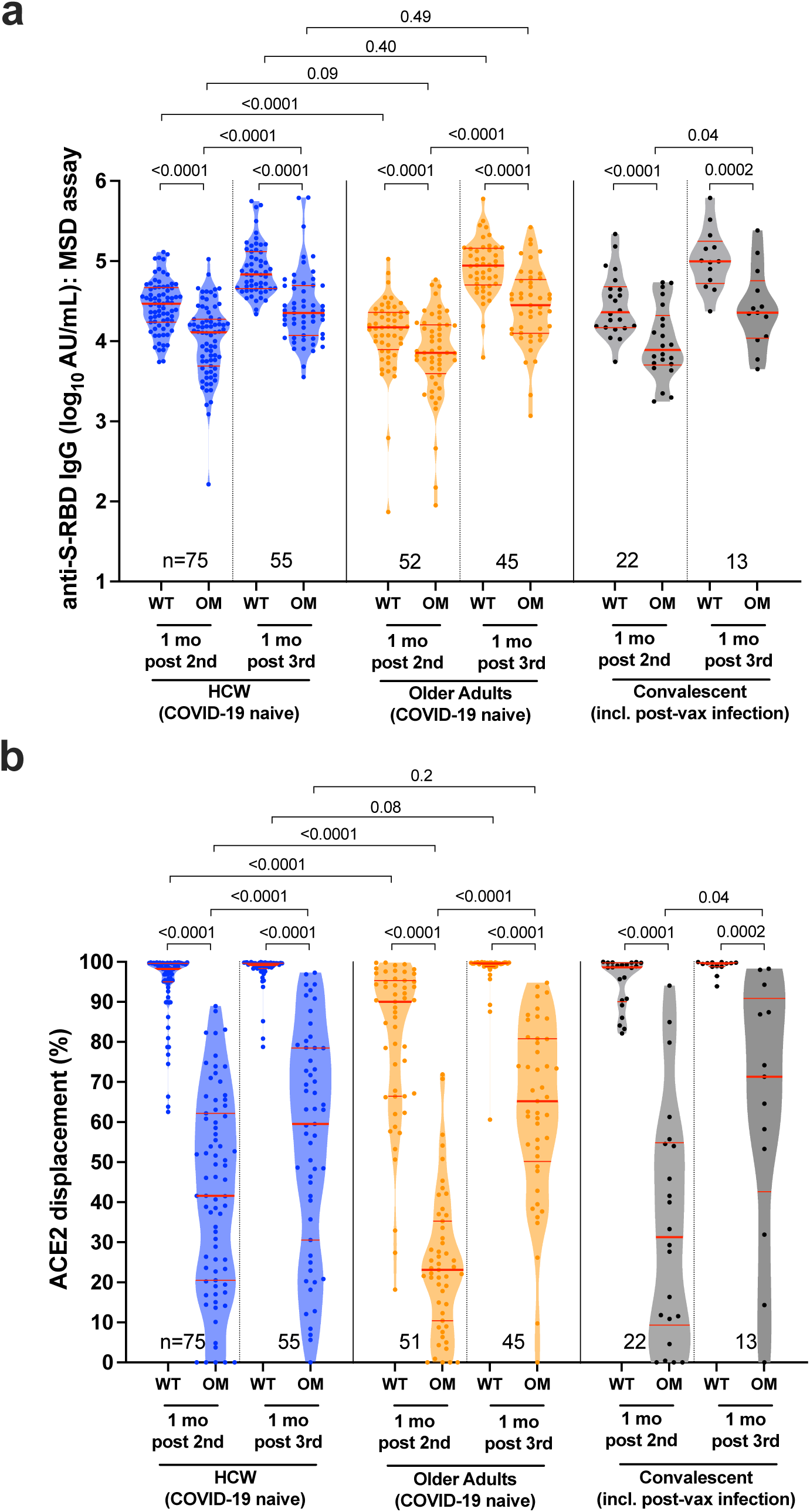
Anti-Omicron IgG binding and ACE2 displacement activities one month after the second and third COVID-19 vaccine doses. *Panel A:* Binding IgG responses in plasma to the wild-type (WT, ancestral Wuhan strain) and Omicron (OM) S-RBD, measured using the Meso Scale Diagnostics (MSD) V-Plex assay, in HCW (blue circles) and older adults (orange circles) who remained COVID-19 naive throughout the study, as well as individuals with prior COVID-19 regardless of infection timing (COVID-19 convalescent; black circles) at one month after the second and third COVID-19 vaccine doses. Participant Ns are shown at the bottom of the plot. A thick horizontal red bar represents the median; thinner horizontal red bars represent the IQR. P-values were computed using the Wilcoxon matched pairs test (for all within-group comparisons) or the Mann-Whitney U-test (for between-group comparisons) and are uncorrected for multiple comparisons. *Panel B:* same as A, but for ACE2 displacement activity, measured using the V-plex SARS-CoV-2 (ACE2) assay, where results are reported in terms of % ACE2 displacement.

We also assessed the ability of plasma to block the interaction between WT and Omicron RBD and the cellular ACE2 receptor, which represents a higher throughput approach to estimate potential virus neutralizing activity (also referred to as a surrogate virus neutralization test [30]). This activity was significantly weaker against Omicron compared to WT RBD after both two and three doses in all groups (all within-group comparisons p≤0.0002; **Figure 3B**), though the discrepancy was most pronounced for older adults after two doses (where median activity against WT was 90% compared to only 23% against Omicron). The third dose universally boosted anti-Omicron activity (all within-group comparisons p<0.05), with, for example, median anti-Omicron activity in older adults rising from 23% after two doses to 66% after three. Consistent with results for binding IgG antibodies, surrogate neutralization of WT RBD was significantly higher in HCW compared to older adults after two doses (p<0.0001), but reached equivalence after three doses (in fact, activities in older adults were slightly higher at this time point; p=0.08). Surrogate neutralization of Omicron RBD followed a similar pattern, with HCW exhibiting significantly higher activity compared to older adults after two doses (p<0.0001), but equivalent levels after three doses (p=0.2). In multivariable analyses, a higher number of chronic health conditions was the strongest correlate of poorer surrogate neutralizing activity against Omicron after three vaccine doses, with each additional condition associated with a ∼6% reduction in this activity (p=0.0046; **Table 3**). Male sex, a longer interval between the first and second doses, and the number of days elapsed since the third dose also correlated with weaker responses after three doses (all p<0.05).

Finally, we assessed plasma neutralizing activity against WT (ancestral USA-WA1/2020 strain) and Omicron using a live virus assay in a subset of 20 HCW and 21 older adults who remained COVID-19 negative throughout the study (**Figure 4**). Neutralizing activity against Omicron was significantly weaker compared to WT following two and three doses in both groups (all p<0.0001). The third dose nevertheless boosted anti-Omicron activity in both groups, where the increase in older adults was particularly pronounced (from a median of BLOQ after the second dose to a median reciprocal dilution of 40 after the third; p<0.0001). Consistent with binding IgG and surrogate neutralization results, anti-Omicron neutralizing activity was significantly lower in older adults compared to HCW after two vaccine doses (p=0.0003) but reached equivalence after the third dose (p=0.79).

**Figure 4:**
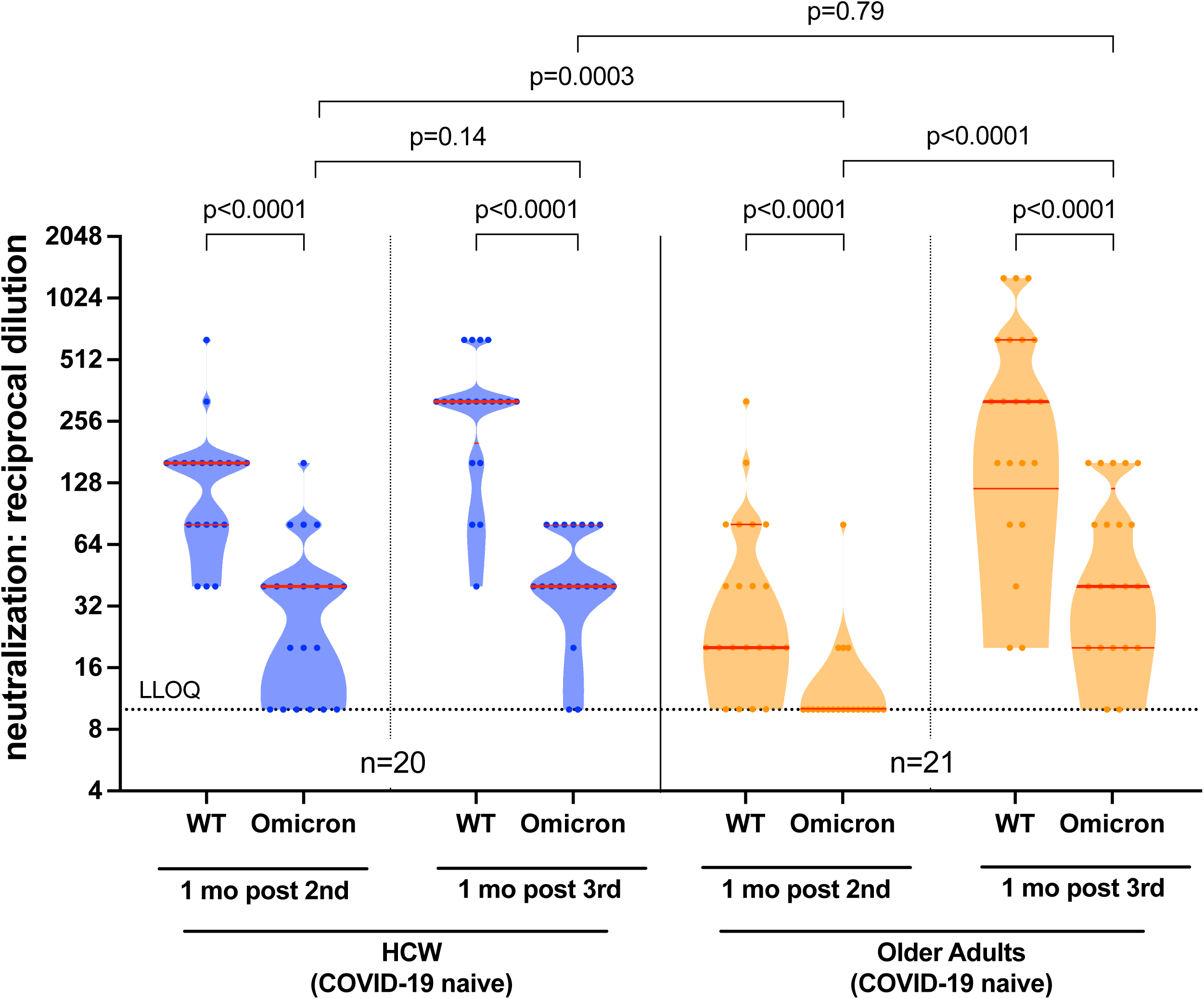
Anti-Omicron neutralization activities one month after the second and third COVID-19 vaccine doses. Neutralization activities, reported as the lowest reciprocal plasma dilution at which neutralization was observed in all wells of a triplicate assay, against the wild-type (WT, ancestral WA1/2020 strain) and Omicron (OM) virus isolates a subset of HCW (blue circles) and older adults (orange circles) who remained COVID-19 naive throughout the study. Participant Ns are shown at the bottom of the plot. A thick horizontal red bar represents the median; thinner horizontal red bars represent the IQR. P-values were computed using the Wilcoxon matched pairs test (for within-group comparisons) or the Mann-Whitney U-test (for between-group comparisons) and are uncorrected for multiple comparisons.

## DISCUSSION

At every time point following two doses of COVID-19 mRNA vaccine, antibody binding and neutralizing activity were significantly weaker in older compared to younger adults. Antibody concentrations were also less durable in older adults, though responses declined substantially in all groups over time (*e*.*g*. by six months after the second dose, neutralizing activities had declined to BLOQ in almost 60% of HCW and >80% of older adults). In multivariable analyses adjusting for sociodemographic, health and vaccine-related variables, a higher number of chronic health conditions remained consistently and independently associated with weaker and less durable binding antibody responses, while a longer interval between first and second doses was consistently associated with higher binding antibody responses after the second dose, as previously reported [24-26]. These findings support public health decisions to provide third doses on or before the six-month mark, with older adults receiving priority.

Third doses of COVID-19 vaccine increased antibody binding and neutralizing function to levels that were significantly higher than those achieved by two doses, where the magnitude of boosting in older adults was particularly prominent. Indeed, antibody binding, surrogate neutralization and live virus neutralization activities in older adults were equivalent to those observed in younger adults after three doses. Consistent with recent evidence [20, 21, 31-37], antibody responses against Omicron were universally weaker than those against the ancestral strain after both two and three vaccine doses; nevertheless, anti-Omicron responses in older adults reached equivalence to those observed in younger adults after three doses. Notably, the number of chronic health conditions persisted as an independent correlate of weaker anti-Omicron responses, even after three doses.

Similar to other reports [27-29], our findings indicate that individuals who have contracted COVID-19 are likely to benefit from vaccination. Compared to naïve participants, convalescent individuals displayed a slower rate of antibody decline, and multivariable analyses demonstrated that binding and neutralization activity was higher in this group at six months after the second dose.

Our study has several limitations. As the precise immune correlates of protection for SARS-CoV-2 transmission and disease severity remain incompletely characterized [38], the implications of our results on individual-level protection from SARS-CoV-2 infection and COVID-19 remain uncertain. We did not investigate T-cell responses, which may play critical roles in protection against severe COVID-19, particularly in the context of variants [39-46]. Our study was not powered to investigate potential differences in immune responses between the two mRNA vaccines [47, 48], nor differences in full vs. half-doses of Spikevax when administered as third doses to individuals ≥70 versus <70 years old, respectively, in Canada. Third dose responses were measured at a single time point, so durability assessments are needed. Nevertheless, results provide additional insight into COVID-19 mRNA vaccine immunogenicity in the elderly and in the context of an extended interval between first and second doses (of up to 112 days).

In conclusion, while the observation of strong binding and neutralizing antibody responses to third COVID-19 vaccine doses in older adults, including to Omicron, are encouraging, it will be important to closely monitor the durability of these responses over time in this population.

## Data Availability

This is an interim report from an ongoing study. Data from this report are available upon reasonable request to the authors in accordance with REB and institutional data sharing requirements. At the end of the study, all data will be deposited in a national database maintained by the COVID-19 Immunity Task Force.

## ACKNOWLEDGEMENTS

We thank the leadership and staff of Providence Health Care, including long-term care and assisted living residences, for their support of this study. We thank the phlebotomists and laboratory staff at St. Paul’s Hospital, the BC Centre for Excellence in HIV/AIDS and Simon Fraser University for assistance. Above all, we thank the participants, without whom this study would not have been possible.

## FUNDING

This work was supported by the Public Health Agency of Canada through a COVID-19 Immunology Task Force COVID-19 “Hot Spots” Award (2020-HQ-000120 to MGR, ZLB, MAB). Additional funding was received from the Canadian Institutes for Health Research (GA2-177713 and the Coronavirus Variants Rapid Response Network (FRN-175622) to MAB), the Canada Foundation for Innovation through Exceptional Opportunities Fund – COVID-19 awards (to MAB, MD, MN, RP, ZLB) and the National Institute of Allergy and Infectious Diseases of the National Institutes of Health (R01AI134229 to RP). MLD and ZLB hold Scholar Awards from the Michael Smith Foundation for Health Research. LYL was supported by an SFU Undergraduate Research Award. GU and FHO hold Ph.D. fellowships from the Sub-Saharan African Network for TB/HIV Research Excellence (SANTHE), a DELTAS Africa Initiative [grant # DEL-15-006]. The DELTAS Africa Initiative is an independent funding scheme of the African Academy of Sciences (AAS)’s Alliance for Accelerating Excellence in Science in Africa (AESA) and supported by the New Partnership for Africa’s Development Planning and Coordinating Agency (NEPAD Agency) with funding from the Wellcome Trust [grant # 107752/Z/15/Z] and the UK government. The views expressed in this publication are those of the authors and not necessarily those of AAS, NEPAD Agency, Wellcome Trust or the UK government.

---

**Table S1:**
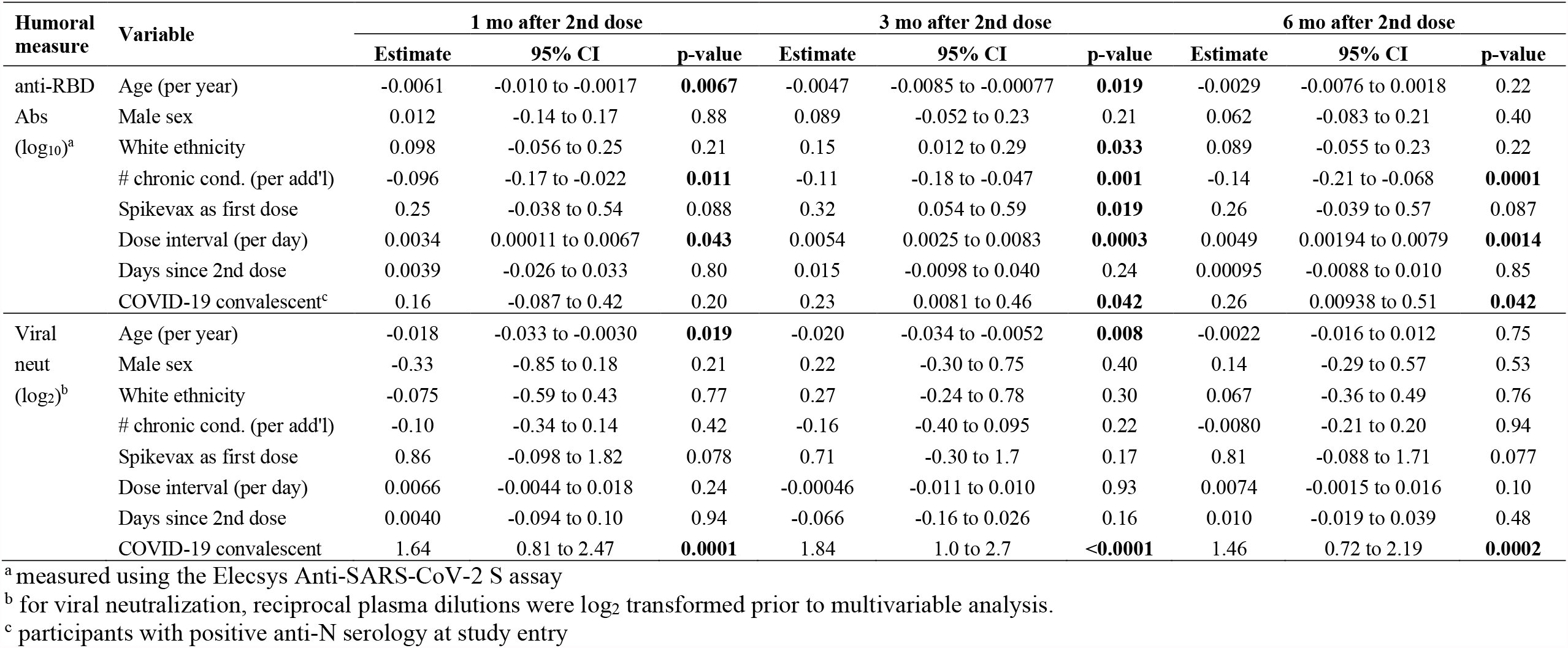
Multivariable analyses of the relationship between sociodemographic, health and vaccine-related variables on immunogenicity measures following two-dose COVID-19 mRNA vaccination.

**Table S2:** Multivariable analyses of the relationship between sociodemographic, health and vaccine-related variables on humoral responses following three-dose COVID-19 mRNA vaccination.

| Humoral measure | Variable | 1 mo after 3rd dose |  |  |
| --- | --- | --- | --- | --- |
|  |  | Estimate | 95% CI | p-value |
| anti-RBD Abs (log<sub>10</sub>)<sup>a</sup> | Age (per year) | 0.0018 | -0.0011 to 0.0048 | 0.22 |
|  | Male sex | 0.0080 | -0.086 to 0.10 | 0.87 |
|  | White ethnicity | 0.0089 | -0.082 to 0.10 | 0.85 |
|  | # chronic conditions (per add'l) | -0.053 | -0.091 to -0.014 | **0.0078** |
|  | Spikevax as third dose (vs. Comirnaty) | 0.12 | 0.030 to 0.21 | **0.0091** |
|  | Interval between 1st and 2nd dose (per day) | -0.00064 | -0.0033 to 0.0020 | 0.63 |
|  | Interval between 2nd and 3rd dose (per day) | 0.00037 | -0.0014 to 0.0022 | 0.69 |
|  | Days since 3rd vaccine dose | 0.0016 | -0.013 to 0.016 | 0.82 |
|  | Prior COVID-19<sup>c</sup> | 0.070 | -0.057 to 0.20 | 0.28 |
| Viral neut. (log<sub>2</sub>)<sup>b</sup> | Age (per year) | 0.022 | 0.0025 to 0.042 | **0.028** |
|  | Male sex | -0.075 | -0.70 to 0.55 | 0.81 |
|  | White ethnicity | -0.28 | -0.88 to 0.33 | 0.37 |
|  | # chronic conditions (per add'l) | -0.17 | -0.43 to 0.081 | 0.18 |
|  | Spikevax as third dose (vs. Comirnaty) | 0.70 | 0.11 to 1.29 | **0.021** |
|  | Interval between 1st and 2nd dose (per day) | 0.012 | -0.0056 to 0.029 | 0.18 |
|  | Interval between 2nd and 3rd dose (per day) | 0.013 | 0.00097 to 0.025 | **0.035** |
|  | Days since 3rd vaccine dose | -0.028 | -0.12 to 0.065 | 0.55 |
|  | Prior COVID-19<sup>c</sup> | 1.23 | 0.39 to 2.06 | **0.0044** |

<sup>a</sup> Measured using the Elecsys Anti-SARS-CoV-2 S assay
<sup>b</sup> Viral neutralization results were log<sub>2</sub>-transformed prior to multivariable analysis
<sup>c</sup> Includes all participants with positive anti-N serology at any time during the study (*i.e.* both pre- and post-vaccine COVID-19 cases)

## Notes

### Competing Interest Statement

The authors have declared no competing interest.

### Author Declarations

This study was approved by the University of British Columbia/Providence Health Care and Simon Fraser University Research Ethics Boards.

### Summary of Updates

Figures 1, 2 and 3 updated to include new data after the second and third vaccine doses; Figure 4 added to show responses agains Omicron strain; Supplemental tables updated.

